# Artificial water fluoridation for dental health improvement: a review and meta-analysis of the evidence and implications of a possible association between water fluoride and IQ

**DOI:** 10.1101/2024.03.08.24303503

**Authors:** Vickie S. Braithwaite, Ruth A. Valentine, Nicholas J. Wareham

## Abstract

**Background:** Globally, dental caries affects 60-90% of schoolchildren. Although artificial water fluoridation improves dental health and reduces dental health inequalities, there is concern that excessive fluoride exposure may lower cognition.

We systematically reviewed and meta-analysed the association between water fluoride and intelligence quotient (IQ).

**Methods:** A literature search of Medline and Web of Science and random-effects meta-analysis comparing mean IQ of children living in low/normal or higher water fluoride areas was investigated. Followed by exploration of possible dose effects among sub-groups, living in moderate (<1.5 ppm) high (1.5-3.0 ppm) or extremely high (>3ppm) water fluoride areas.

**Results:** Twenty-three observational studies (n=9539 children) were included. Overall, the higher water fluoride group had a lower mean IQ compared with the low water fluoride group (standardised mean difference (95% confidence interval): -0.43 (-0.63 to -0.24) p<0.0001, I^2^=94.2% p<0.0001). Sub-group analysis showed no association between water fluoride and mean IQ in studies of moderate fluoride concentrations (moderate: 0.04 (-0.08 to 0.15) p=0.53, I2=0.0% p=0.68). Mean IQ was lower in the higher water fluoride groups (high: - 0.52 (-0.92 to -0.12) p=0.01, I^2^=96.2% p<0.00001, extremely high: -0.60 (-0.87 to -0.33), p<0.0001, I^2^=84.6% p<0.0001).

**Conclusions:** At moderate levels (<1.5 ppm) there was no statistical or clinically meaningful association between water fluoride and IQ. This suggests that populations living in these areas could benefit from artificial water fluoridation without experiencing neurotoxicity. An association between lower mean IQ and high water fluoride observed suggests a need to prioritise removal of excess fluoride from drinking water in these regions.

**Key Message (3-5 bullet points in complete sentence):** At low concentrations of water fluoride (<1.5 ppm) there was no detectable association between water fluoride and intelligence quotient (IQ);

At high levels of water fluoride concentrations (>1.5 ppm) there was an inverse association between increasing fluoride concentration and decreasing mean IQ;

In countries in which fluoridation of water is considered but kept within safe concentrations, the available evidence suggests that fluoridation has demonstrable benefits on public dental health without any clinically significant effect on IQ.

In countries with excessive groundwater fluoride concentrations there is an imperative to promote water de-fluoridation schemes to protect the public from skeletal and dental fluorosis and from possible neurotoxicity.

## Background

Poor dental health in children is of major public health concern. Globally, dental caries are estimated to affect 60-90% of schoolchildren and the vast majority of adults (1) and is the most common non-communicable disease (2). As well as the direct medical costs, ∼US$300 billion spent on dental caries globally per year (2), there are substantial indirect medical and societal costs associated with poor dental health in children include taking time off school, dental pain, decreased attainment in school and loss of parental earnings (3).

Tooth decay is caused by the production of acid from the breakdown of fermentable carbohydrates by bacteria in biofilm on tooth surfaces. This process is exacerbated by dietary factors such as consumption of sugar, acid and carbonated beverages (4) and is strongly associated with deprivation. For example, tooth decay in children living in the most deprived counties in England is almost twice as high compared with children living in the least deprived counties (30% and 16%, respectively)(**Figure 1**).

**Figure 1.**
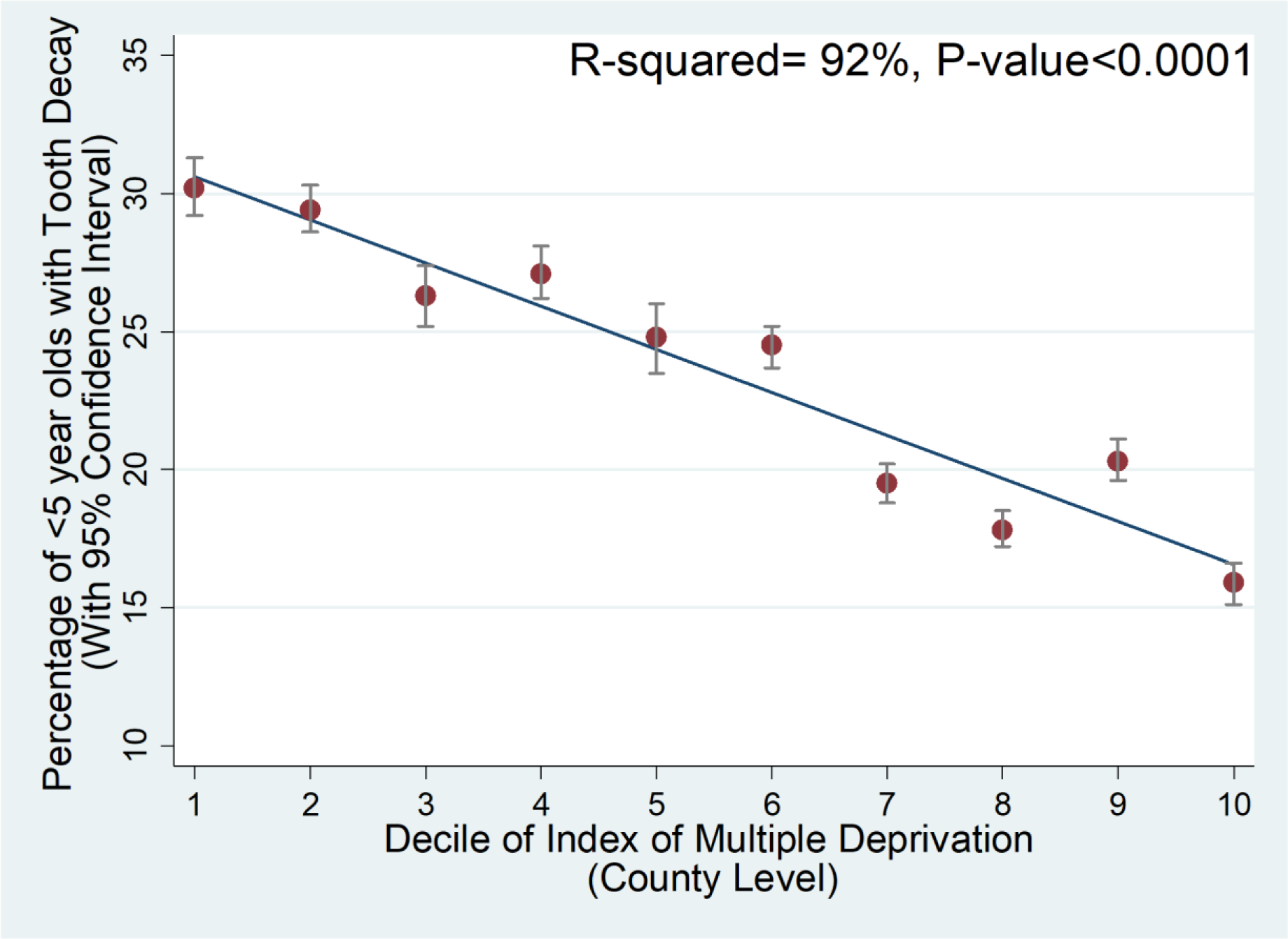
The association between index of multiple deprivation and percentage of children <5 years of age with tooth decay in England, UK. There was a negative association between index of multiple deprivation (county and unitary authority level data) and % of <5 year olds with tooth decay in England (P<0.0001). 92% of the variation in tooth decay was explained by index of multiple of deprivation. 1 is the most deprived decile and 10 is the least deprived decile. 95% confidence intervals were included in the figure as the error bars. Data to produce this graph was obtained from Public Health England using 2017 data (1). **NB.** Y-axis starts at 10% for visual clarity.

Fluoride exposure can protect against tooth decay (5). Calcium-hydroxyapatite is the major inorganic component of enamel as well as other hard tissues such as dentine and bone (4). When exposed to acid, calcium-hydroxyapatite dissolves resulting in tooth-softening and decay (4). Fluoride can reduce demineralisation and enhance remineralisation of enamel by displacing calcium and forming fluorine-hydroxyapatite; a compound that is less soluble than calcium-hydroxyapatite (4, 6).

The main methods of ensuring topical exposure of fluoride in many high income countries is through tooth-brushing with fluoride-containing toothpaste and through applying fluoride-varnishes to teeth by dental professionals. Despite these interventions, tooth decay remains a major problem. Many health organisations including Public Health England (PHE), the Centre for Disease and Control and Prevention and the WHO promote artificial water fluoridation as a solution to the problem of poor dental health in regions with low levels of naturally occurring fluoride (1, 7).

The potential benefits of artificial water fluoridation have been long known. Grand Rapids, a city in Michigan, USA was the first city in the world to fluoridate their water supply in the 1940s and to show a dramatic reduction in dental caries following this intervention (8). With artificial water fluoridation, the community water supply is supplemented with a silicofluoride compound. 1.0 ppm (parts per million) of fluoride in water is considered optimal for dental health (9) and 1.5 ppm is considered the maximum upper-limit for overall health (10).

Water fluoridation is an effective primary prevention measure and has been shown to reduce tooth decay in deciduous and permanent teeth by ∼25-35% and to reduce tooth decay-related paediatric hospital admissions by 55% (11-13). Importantly, water fluoridation is a low-agency intervention and as such would be expected to improve overall dental health and can reduce dental health inequalities (14). It has been estimated that ∼370 million people worldwide, in 27 countries are supplied with artificially fluoridated water (15), however, in the UK only 10% of the population has access to naturally (>0.5 ppm) or artificially fluoridated water (16) and this pattern is mirrored in many other European countries

Despite the effectiveness of artificial water fluoridation in improving dental health, there are concerns about the safety of fluoride exposure on health and the ethics of the intervention (17, 18). Fluoride occurs naturally in groundwater but the concentration varies depending on underlying geology (19). Naturally occurring concentrations of groundwater fluoride is typically low across Europe (<0.5 ppm) whereas concentrations >25 ppm have been reported in India, China and Africa (19) where cases of dental and skeletal fluorosis are also seen. Dental and skeletal fluorosis are defined by the incorporation of excessive amounts of fluoride into calcified tissues. Mild dental fluorosis results in cosmetic discolouration of teeth with no other side effects (20), whereas severe dental fluorosis results in dark-brown mottling of teeth, porosity of enamel and increased risk of tooth decay (21). Skeletal fluorosis can lead to an increased fracture risk (22).

However, beyond the skeletal adverse effects of high fluoride exposure, there is growing concern that fluoride exposure in early childhood may act as a neurotoxin and impair cognitive ability as measured by intelligence quotient (IQ) (17).

The aims of this study were to review and meta-analyse the published literature investigating a possible association between water fluoride concentration and IQ and to discuss these findings in the broader context of artificial water fluoridation.

## Methods

### Search Strategy of the Literature

A literature search was performed of Medline and Web of Science on the 08-January-2020 and inclusion and exclusion criteria were developed together with the study aims prior to performing the literature search (**Table 1 and 2**). The search, literature review, assessment of study quality and data abstraction was performed by a single reviewer.

**Table 1.**
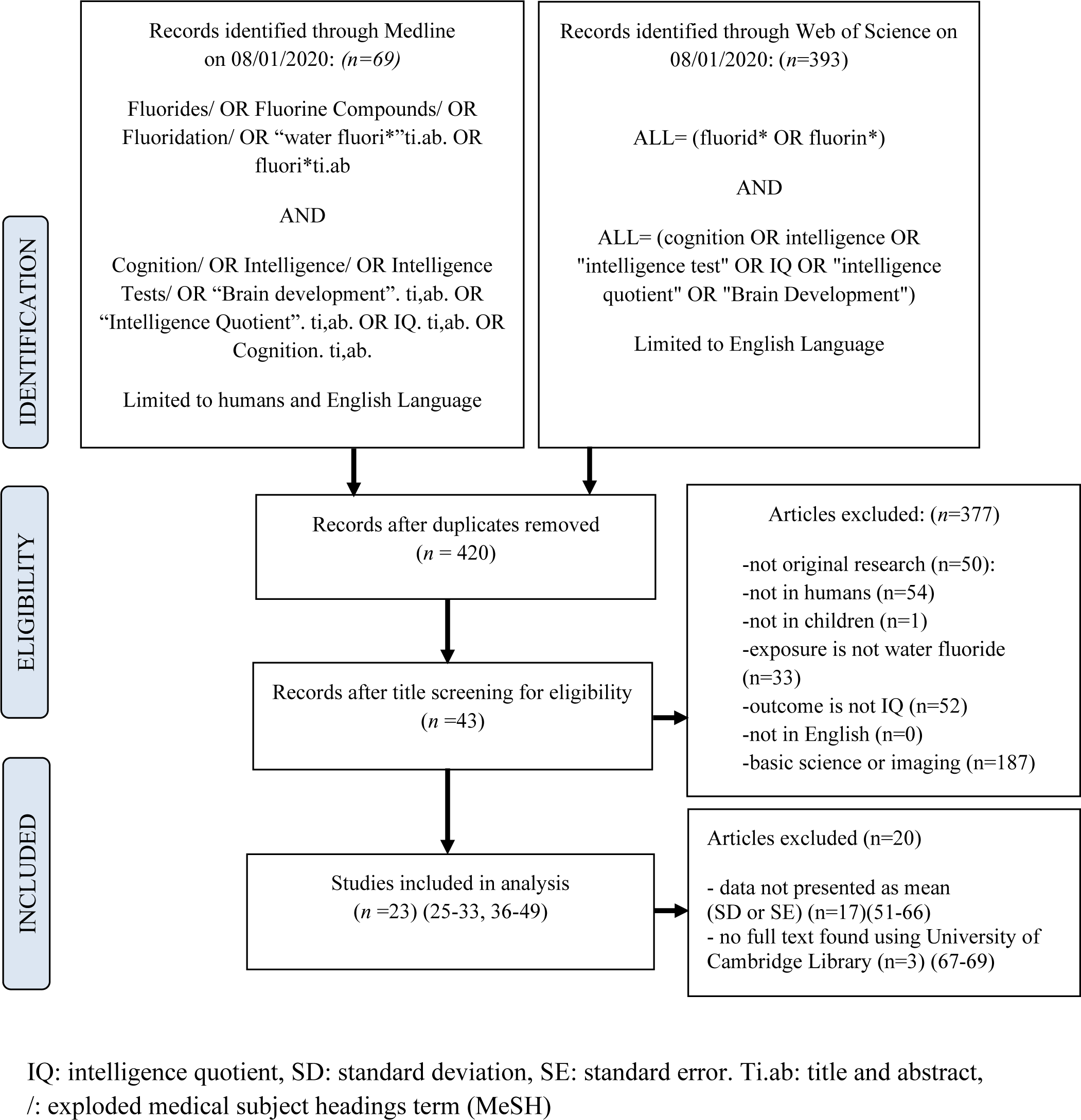
PRISMA flow-diagram of search strategy, search results and studies included in the final meta-analysis.

### Assessment of Study Quality

All studies identified in the literature search were observational studies. Study quality was assessed using the Newcastle-Ottawa scale for cohort studies (23) and adapted for cross-sectional studies (24). Studies were scored in three domains: selection, comparability and outcome and were scored out of a maximum of 10 points. Studies with scores >7 were classed as high quality, 5-7 as good and 0-4 as poor.

### Statistical Analysis

A random effects meta-analysis model was selected *a priori* based on the anticipated heterogeneity of studies. The meta-analysis was performed using sample size, mean IQ and standard deviation (or standard error, subsequently converted to standard deviation) (25, 26) of studies assessing the standardised mean difference (SMD) in IQ in children exposed to low/normal concentrations of water fluoride compared with children exposed to higher concentrations of water fluoride. For studies that presented total IQ and a breakdown of IQ (e.g. verbal and performance), total IQ score was used (27, 28). Where studies presented more than two comparator groups i.e. low, moderate and high water fluoride concentrations the moderate groups was selected to compare with the low fluoride group and the high group was excluded; the low to moderate comparison was deemed to be more informative than low to high comparison. (29-33).

The meta-analysis was performed using the STATA programme “metan” and was presented as SMD and 95% confidence interval (CI) and I^2^ to indicate the extent of study heterogeneity (34). Studies were weighted using study standard deviation (σ) and between study standard deviation (τ; as a measure of study heterogeneity) using the following formula: weight = 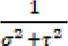. Publication bias was assessed using a funnel plot and Egger’s test using the programmes “metafunnel” and “metabias” respectively (35).

Subgroup analysis was performed based on mean fluoride concentration. Studies were categorised as moderate: <1.5 ppm, high: 1.5-3.0 ppm, extremely high: >3.0 ppm or unknown.

Sensitivity analysis was performed to assess the effect of outliers on the meta-analysis.

## Results

### Study Characteristics

Twenty-three studies from five different countries were identified; three were prospective cohort studies from New Zealand (36) and Canada (27, 37) and 20 were cross-sectional studies from China, India and Iran (**Table 3**)(25, 26, 28-33, 38-49). Data from 9539 children were used and participants were aged 3-14 years. Nineteen of 20 cross-sectional studies compared the IQ of children living in regions with naturally high or low concentrations of groundwater fluoride. The three prospective cohort studies compared the IQ of children living in regions with naturally low concentrations of groundwater fluoride with or without artificial water fluoridation programmes. The mean fluoride concentration in the reference group was ≤1.5 ppm in all studies (range 0.1-1.5 ppm) with the exception of one study with a mean of 2.0 ppm (26). Two studies did not specify fluoride concentration (**Table 3**)(41, 45). The three prospective cohort studies and one cross-sectional study compared their reference group with a moderate fluoride group (<1.5 ppm) and the remaining 17 cross-sectional studies compared their reference groups with high (1.5-3.0 ppm, n=11 studies) or extremely high (>3.0 ppm, n=6 studies) fluoride groups. Study quality ranged from poor to high quality (2-9) with a median score of 5 (interquartile range: 5-6) (**Table 3**).

**Table 2.** Inclusion and exclusion criteria.

| **Inclusion Criteria** | **Exclusion Criteria** |
| --- | --- |
| Article is in humans | Editorials, reviews, conference abstracts |
| Article looks at water fluoride exposure | Full text not available |
| Article presents mean and standard deviation/error of intelligence quotient in low/normal and higher concentrations of water fluoride concentration | Not available in English |
|  | Not in children |

**Table 3.**
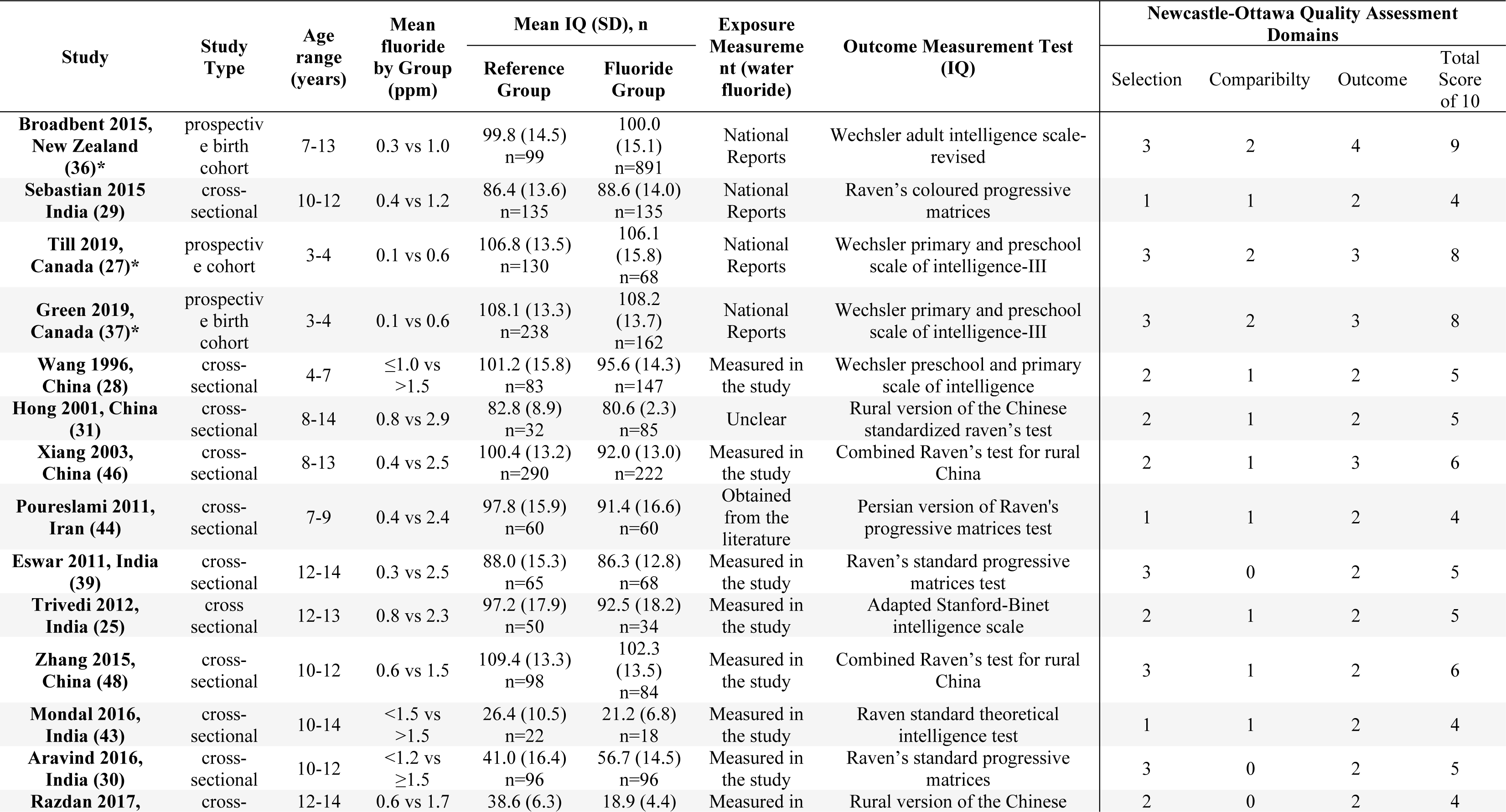
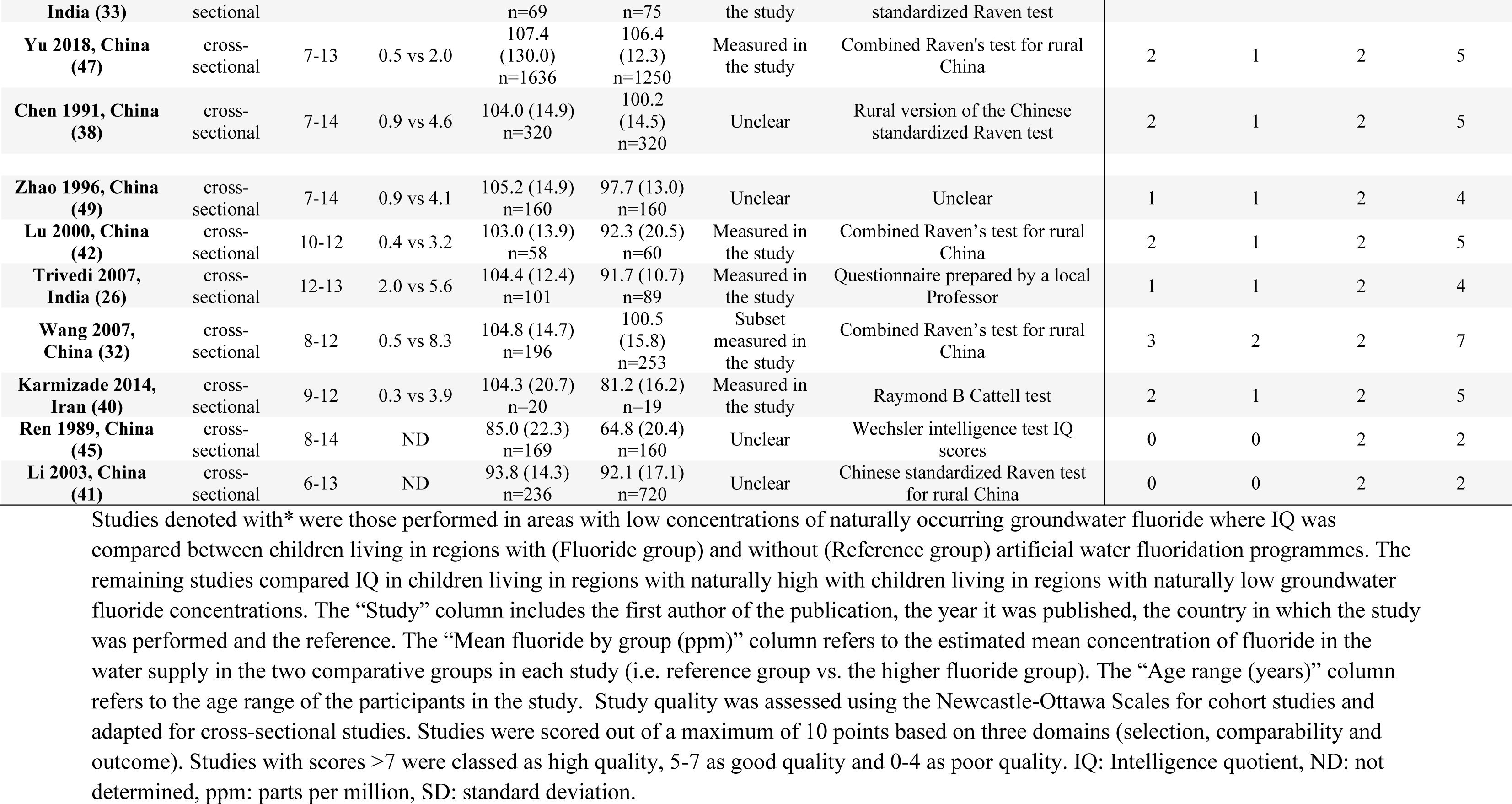
Characteristics and quality of the studies found through the literature search.

### Meta-analysis

The meta-analysis estimated a pooled effect of a lower IQ in the high fluoride groups compared with the low fluoride groups; standardised mean difference (95% CI) of -0.43 (- 0.63 to -0.24) p<0.0001 with significant study heterogeneity (I^2^=94.2% p<0.0001) (**Figure 2**).

**Figure 2.**
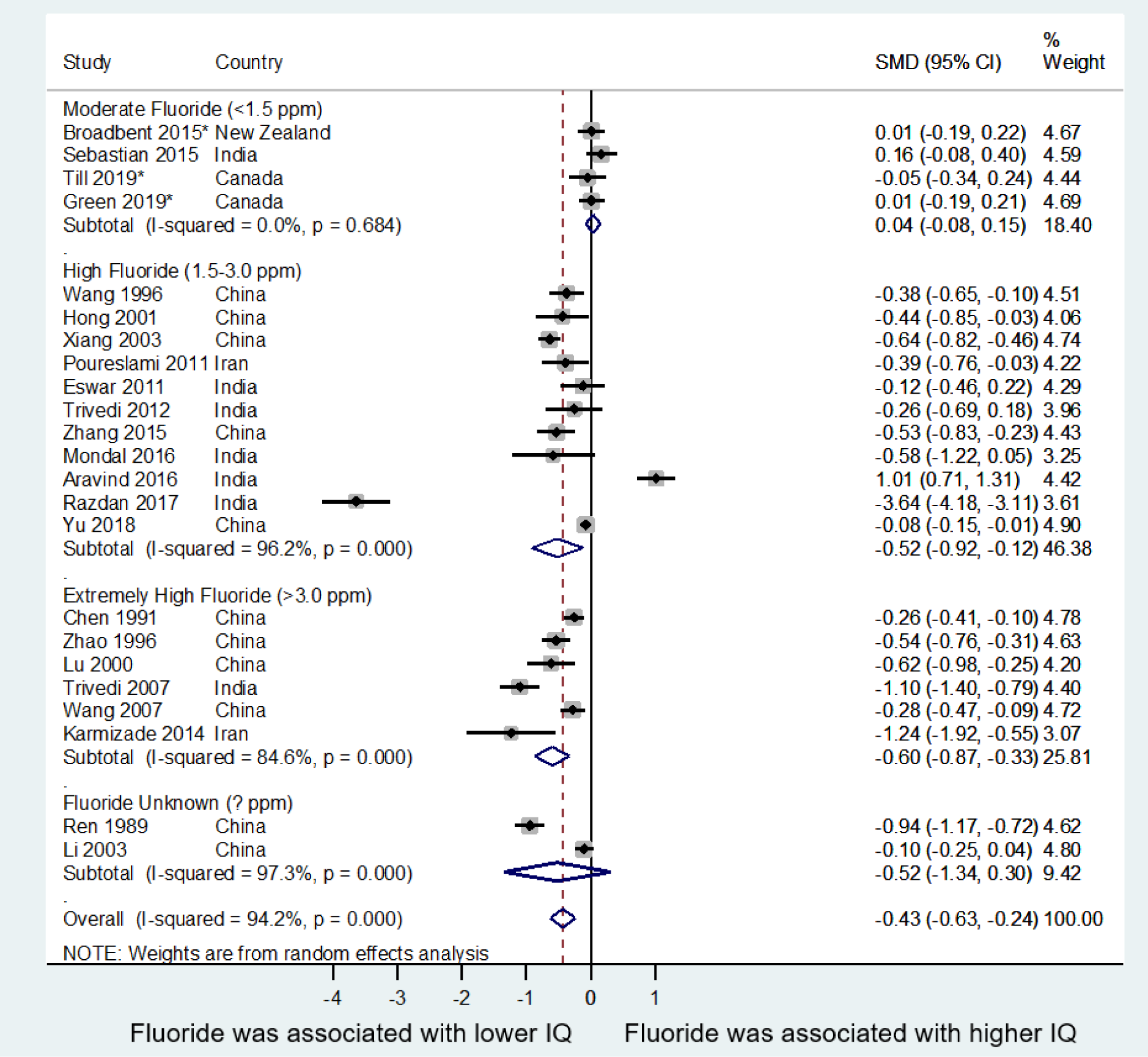
Random effects meta-analysis of the association between water fluoride and intelligence quotient (IQ) Random effects meta-analysis of the standardised mean difference (SMD) of intelligence quotient (IQ) in children from the reference group compared with the higher fluoride group. *denotes studies conducted in countries with low concentrations of naturally occurring groundwater fluoride with and without artificial water fluoridation programmes. The remaining studies compared IQ in regions with moderate or high with low concentrations of naturally occurring groundwater fluoride. I^-^squared was a measure of study heterogeneity. Subgroup analysis was performed by categorising studies based on the mean fluoride concentration of their higher fluoride group: moderate (<1.5 ppm, n=4), high (1.5-3.0 ppm, n=11), extremely high (>3.0 ppm, n=6) and unknown (n=2). The concentration of water fluoride in the reference group was <1.5 ppm in all studies (range 0.1-1.5 ppm) with the exception of three studies; one study that had a mean concentration of 2 ppm (26) and two that did not specify water fluoride concentration (41, 45). Overall, mean IQ was significantly lower in the higher fluoride group compared to the reference (SMD (95% CI): -0.43 (-0.63 to -0.24), p<0.0001) and there was substantial study heterogeneity (I-squared: 94.2%, p<0.0001). Subgroup analysis indicated no significant difference in IQ in the moderate fluoride group compared with the reference (0.04 (-0.08 to 0.15), p=0.53) and no study heterogeneity was detected (I-squared: 0.0%, p=0.68). IQ was significantly lower in the high fluoride (-0.52 (-0.92 to -0.12), p=0.01) and extremely high fluoride groups (-0.60 (-0.87 to - 0.33), p<0.0001) compared to the reference and there was substantial heterogeneity (I-squared: 96.2% and 86.6%, p<0.0001 respectively). There was no significant difference in mean IQ in the subgroup of studies with unknown fluoride concentrations (-0.52 (-1.34 to 0.30), p=0.22). CI: confidence interval, IQ: intelligence quotient, ppm: parts per million, SMD: standardised mean difference.

Subgroup analysis of studies grouped by the fluoride concentration showed no association between water fluoride concentration and IQ in the moderate fluoride group compared with the reference (<1.5 ppm: 0.04 (-0.08 to 0.15) p=0.53) and no study heterogeneity was detected (I^2^=0.0% p=0.68). IQ was significantly lower in studies with high or extremely high water fluoride concentrations compared with the reference (1.5-3.0 ppm: -0.52 (-0.92 to - 0.12) p=0.01 and >3.0 ppm: -0.60 (-0.87 to -0.33) p<0.0001) (**Figure 2**) and study heterogeneity was significant (I^2^=96.2% and 84.6% p<0.0001 respectively).

### Bias

Assymmetry of the funnel plot showed evidence of publication bias (Egger’s test p=0.055)(**Figure 3**). In particular, there was an absence of small studies showing a null or a positive association between water fluoride concentrations and IQ. The funnel plot indicated a clear study outlier (33). When this study was excluded the pooled effect size decreased to - 0.31 (-0.46, -0.14) p<0.0001 and I^2^ decreased to 91% p<0.0001 (data not shown).

**Figure 3.**
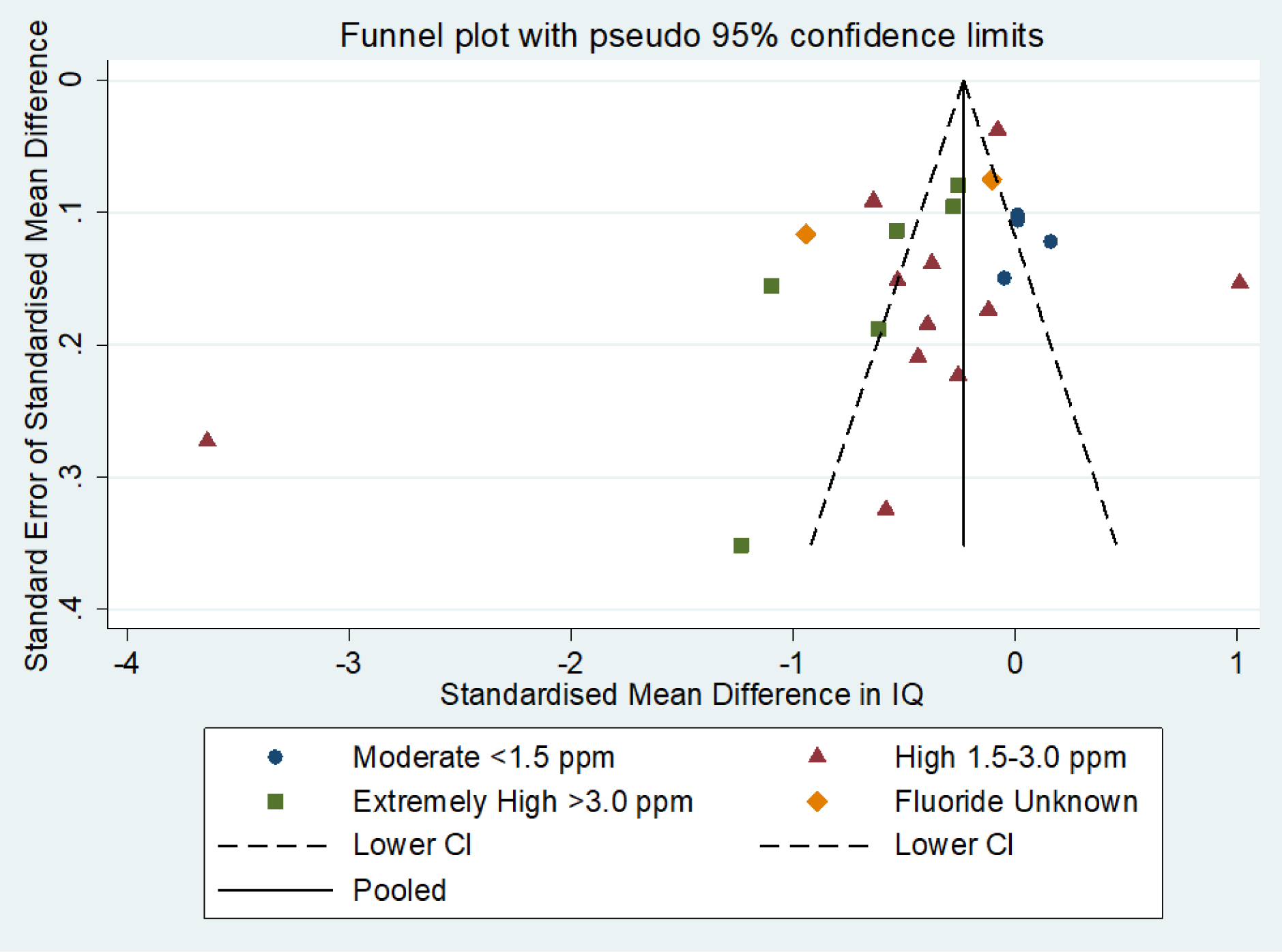
Assessment of possible publication bias with a funnel plot and Egger’s test. Studies were grouped by the mean fluoride concentration of their fluoride group: moderate (blue, <1.5 ppm, n=4), high (red, 1.5-3.0 ppm, n=11), extremely high (green, >3.0 ppm, n=6) and unknown (orange, n=2). The Funnel plot indicated a clear outlier study conducted in a region of India with naturally high groundwater fluoride (33). The plot suggested possible publication bias as there was asymmetry in the graph; particularly a lack of studies with higher standard error (i.e. smaller studies) that show a null or a positive association between fluoride concentration and intelligence quotient. Egger’s test for bias was p=0.055. CI: confidence interval, IQ: Intelligence quotient, Pooled: meta-analysis pooled-effect size, ppm: parts per million.

## Discussion

The random-effects meta-analysis indicated that, overall, there was a negative statistical association between water fluoride and mean IQ (SMD (95% CI): -0.43 (-0.63 to -0.24), p<0.0001). This finding was based on 23 studies consisting of 9,539 children.

However, at moderate concentrations of water fluoride that were below the considered safe- upper limit (<1.5 ppm), there was no association between water fluoride and mean IQ (0.04 (- 0.08, 0.15), p=0.53). This finding was based on four studies (*n=*1858 children), three of which were cohort studies that compared IQ in regions with or without artificial water fluoridation programmes in countries with low concentrations of naturally occurring groundwater fluoride (New Zealand and Canada).

In the studies with high (1.5-3.0 ppm) or extremely high (>3.0 ppm) concentrations of water fluoride, there was a significant, negative association between water fluoride and IQ (high: - 0.52 (-0.92, -0.12), p=0.01 and extremely high: -0.60 (-0.87, -0.33), p<0.0001). These finding were based on 17 cross-sectional studies (n=6396 children) that compared IQ in regions with high or low concentrations of naturally occurring groundwater fluoride (China, India and Iran).

While there was evidence to suggest an association between high or extremely high concentrations of naturally occurring water fluoride (≥1.5 ppm) and a lower IQ, there was no statistical evidence of an association between artificial water fluoridation (<1.5 ppm) and IQ in regions with naturally low concentrations of groundwater fluoride.

This is the first meta-analysis investigating the association between water fluoride and IQ to include the three cohort studies comparing IQ in regions with or without artificial water fluoridation programmes (27, 36, 37). Previous meta-analyses only included studies from regions with naturally high concentrations of groundwater fluoride (50). In addition, this meta-analysis attempted to investigate whether the association between water fluoride and IQ depended on fluoride concentration (i.e. moderate, high and extremely high). Crucially, this is the first meta-analysis to suggest that there is no association between moderate concentrations of water fluoride and IQ. Our results are consistent with other meta-analyses suggesting an inverse association between fluoride and IQ when concentrations are high (27, 36, 37).

The studies included in this meta-analysis ranged in quality. The three cohort studies were assessed as being of high quality and the remaining cross-sectional studies were assessed as being of poor or good quality. There was substantial variation in study design, most of the studies were susceptible to bias and confounding and causality was not established. In addition, most of the studies were semi-ecological by design and did not assess individual fluoride exposure through total daily fluoride intake measurements or through biological markers of individual fluoride exposure (e.g. fluoride in urine, blood, hair).

A further weakness was that the subgroup of studies with moderate fluoride concentration (<1.5 ppm) comprised of 1858 participants only. This sample size may have been underpowered to detect a small difference in mean IQ where group differences in fluoride concentrations was also small.

## Conclusion

This meta-analysis suggests that water fluoridation at moderate concentrations (<1.5 ppm) was not associated with lower IQ in regions with low concentrations of naturally occurring groundwater fluoride. This suggests that artificial fluoridation in these regions may offer substantial public dental health benefit without any neurotoxic effects although this finding was not based on individual level fluoride exposure. However, there was evidence of an association between water fluoride concentrations and lower IQ in regions with high concentrations of naturally occurring groundwater fluoride (≥1.5 ppm). In such countries, there is an imperative to promote water de-fluoridation schemes to protect the public from skeletal and dental fluorosis and from possible neurotoxicity.

## Data Availability

All data produced are available online through a review of the literature

